# Telemedicine in Surgical Care During COVID-19 in LMICs: A Structured Review of Implementation and Impact

**DOI:** 10.1101/2025.05.25.25328225

**Authors:** Tiffany Truong

## Abstract

**Background:** The COVID-19 pandemic severely disrupted surgical care worldwide, with particularly acute effects in low- and middle-income countries (LMICs). Telemedicine (TM) was rapidly adopted to mitigate these disruptions, but evidence on its role in surgical care in LMIC settings remains limited.

**Objective:** To review the implementation, impact, and challenges of telemedicine in surgical care across LMICs during the COVID-19 pandemic.

**Methods:** Following PRISMA 2020 guidelines, we searched PubMed, Embase, Web of Science, and Ovid for peer-reviewed studies from December 2019 to July 2022. Studies assessing TM interventions in any surgical specialty within LMICs were included. Data extraction focused on TM platform types, patient and provider outcomes, feasibility, and policy implications. Quality was appraised using the Joanna Briggs Institute tool for observational studies.

**Results:** Thirteen studies across six surgical specialties and 4,155 patients were included. TM was used for follow-up (46%), consultation (38%), and remote treatment (23%), with high patient satisfaction (mean ≥ 85%), improved access, and cost savings reported. Four studies noted positive clinical outcomes (e.g., reduced complications, optimized medication). Barriers included connectivity issues, regulatory gaps, lack of physical examination capability, and infrastructure inequities.

**Conclusions:** TM provided feasible, safe, and effective surgical support during the pandemic in LMICs, particularly in rural settings. However, long-term sustainability requires investment in digital infrastructure, standardized protocols, and data privacy regulation. TM should be integrated into national surgical planning beyond COVID-19.

**Taxonomy in Telemedicine:** Several terms have been used to indicate remote health services, therefore it is essential to differentiate them, understand their scope and their relation to each other [1].

- **E-health:** The term includes all types of secure use of information and communication technology (ICT) related to health, for example, applications and websites for health promotion, education, screening, research, assessment, and virtual video-chat sessions [2].
- **Telehealth:** Compared to e-health, telehealth is limited to healthcare over a distance. It is more extensive than telemedicine and incorporates educational activities related to patients and providers, public health intervention, and health administration [3].
- **Telemedicine (TM):** A subgroup of telehealth that focuses only on the curative aspect. It can be divided into medical specialties such as dermatology (tele-dermatology), psychiatry (telepsychiatry), and radiology (teleradiology).
- **Telecare:** A subgroup of telehealth that focuses on the preventive aspect. It provides automated monitoring of behavior changes over time.
- **mHealth:** Mobile technologies that can have a spectrum of purposes in the delivery of care.

Figure 1:
the scope and relationship between the different telehealth related terms.

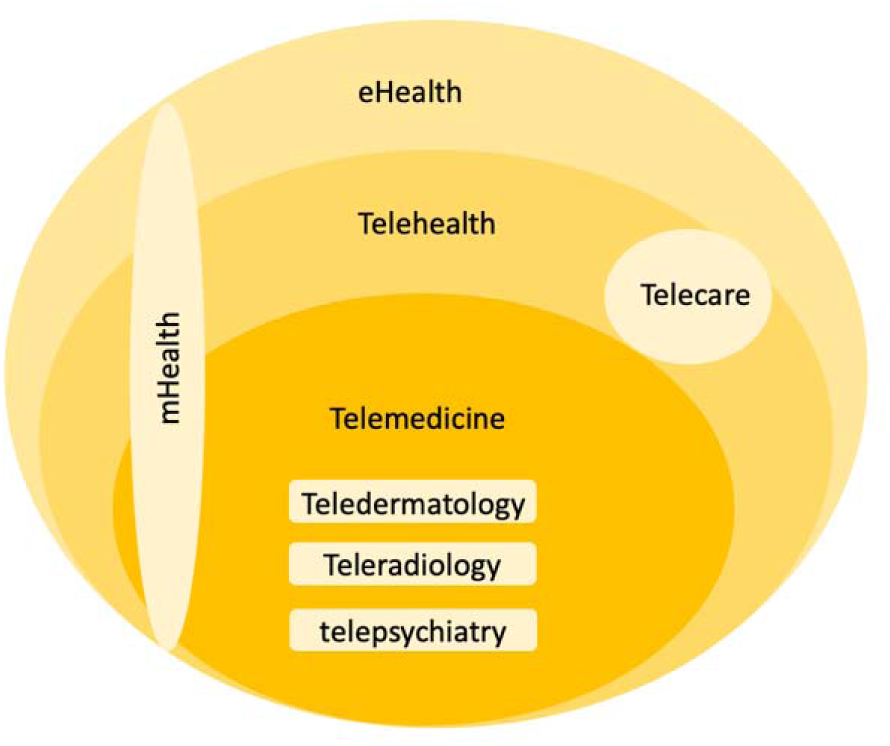

## 1. Introduction

Surgical care is essential globally, with 310 million major surgeries performed to treat numerous life-threatening conditions [4, 5]. While 28% of the overall global burden of disease is surgical, it disproportionately affects developing countries [6]. With urbanization and industrialization, there has been an increase in work and vehicle-related injuries, increasing the need for trauma and surgical care [7]. Moreover, the shift from infectious diseases to chronic and non-communicable diseases (NCDs) amenable to surgical interventions has increased reliance on these services [8].

The COVID-19 pandemic further compounded the existing access barriers. The lockdowns and unprecedented movement restrictions led to drastic reductions in surgical capacity due to the redirection of medical attention to contain and treat patients infected with COVID-19. The limited medical resources and protective equipment reinforced this rationale. Therefore, the surgical field was heavily disrupted by the near-complete global cancellation of surgical services [9]. Delayed electives surgeries can create serious backlogs and worsens health outcomes up to 50% with just a six-months delay [10].

To mitigate the disruptions, telemedicine (TM) was implemented - defined as the practice of medicine by health professionals using telecommunication technologies such as video calls, texts, online messaging, and emails (AAFP). Governments encouraged its usage to reduce footprints in medical facilities (Mahajan et al, 2020). Existing systematic reviews conducted in high-income countries (HICs) have shown that TM improves health access and can be cost-effective [11]. However, evidence in LMICs remains limited. HIC-centric findings may not be generalizable due to differing infrastructure, workforce, and regulatory contexts. As TM became a policy priority in LMICs to reduce the impact of COVID-19 restrictions on health access, an evidence synthesis is critical. This structured review aims to do so by evaluating TM’s implementation, impact and challenges in surgery in LMICs during COVID-19. It aims to inform policy, identify gaps and propose future directions for sustainable digital health integration post-pandemic.

**Figure:**
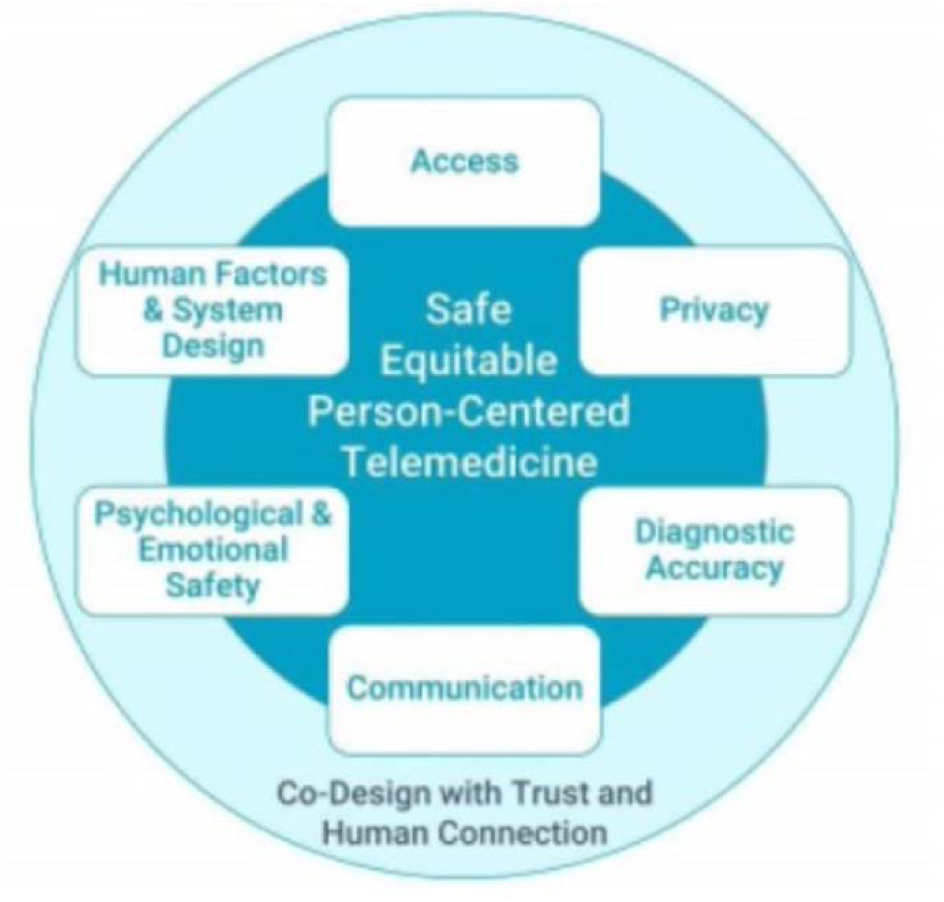
Theoretical framework for patient-centered telemedicine [12]

The theoretical framework from the Institute for Healthcare Improvement (IHI) on person-centered telemedicine was used to evaluate the impact [12]. The core components which include access, privacy, human factors/system design, psychological and emotional safety, communication, and diagnostic accuracy, will be used to assess the TM’s utility and limitations.

## 2. Methods

A structured review was conducted according to the PRISMA 2020 protocol to synthesize the implementation, impact, and challenges of telemedicine in surgical care across LMICs during the COVID-19 pandemic Peer-reviewed studies were selected from Embase (OVID), PubMed, and Web of Science from December 31st, 2019, to July 28^th^, 2022, the peak of the Pandemic.

A search strategy was formulated with the consultation of an experienced librarian. It was developed based on the PICOS framework to address all keywords of the research question in the data collection. A combination of keywords (“telemedicine”, “surgery”, “COVID-19”, “LMIC”) with their synonyms and the database sub-heading terms were used to maximize the collection coverage. The list of countries to be included follows the World Bank classification system for low and middle-income countries [13]. The different concepts were merged using Boolean operators “OR” and “AND”. Systematic reviews were excluded. The detailed search strategy for each database is detailed in Appendix 1.

### a. Study selection and eligibility criteria

Due to the context of dissertation, the author was the only individual to process the screening, extraction and synthesis. Reports that are peer-reviewed that evaluate or assess the impact of TM in any surgical specialty in LMICs were included. The time frame of interest is during the COVID-19 pandemic, thus studies published before and not COVID-related were excluded. The PICOS framework was used as the reference for data extraction as well as eligibility criteria.

### b. Quality appraisal

The quality assessment of included studies was done using the Joanna Briggs Institute (JBI) Critical Appraisal Tool for quantitative descriptive studies such as cross-sectional, retrospective and prospective studies [14]. This tool was used to evaluate the quality and bias of the studies in terms of their methods of design, sampling, data, and results collection. Only studies that have satisfied at least 80% of the criteria listed.

### c. Data extraction and analysis

Data from the 13 included studies were charted in a Microsoft Excel sheet. The information extracted included publication year, country, setting (rural, urban), aim, study design, participants’ characteristics, surgical specialty, the telemedicine platforms, and the key findings. The types of study outcomes included data from both patients and physicians regarding the feasibility and effectiveness of telemedicine. Given the nature of the papers included, a meta-analysis was not performed. A structured and narrative analysis of the impact of TM platforms was conducted instead.

## 3. Results

### a. Eligible studies

Data extraction generated OVID (91) and PubMed (175) and Web of Science (32). The total reports to be screened were 141 and 13 studies satisfied the inclusion criteria.

### b. Studies characteristics

13 studies were included in the systematic review, covering six surgical specialties represented: neurosurgery (6), general surgery (3), urology (1), oncology (1), pediatrics (1), and ENT (1). A summary is presented in Table 1. All studies were conducted in LMICs, as classified by the World Bank. Of the included studies, 76% (10/13) originated from Asia, with seven conducted in China. One study was from Egypt and one from Brazil. validated clinical scales were used such as the NOSE (Nasal obstruction and symptom evaluation), Self-Rating Depression Scale (SDS), the Self-Rating Anxiety Scale (SAS), Yale-Brown Obsessive-Compulsive Scale, (Y-BOCS) (Tiwari et al., 2021; Zhang, Huang et al., 2020; Lin et al., 2022).

**Table 1:** Characteristics of included studies.

| Author and Date | Title | Study Aim | Journal | Country | Funding Source | Transparency |
| --- | --- | --- | --- | --- | --- | --- |
| **Qiu et al, 2020** | Impact of covid to Patients with IBD from Hubei province (the epicenter of COVID-19) and Guangdong province (a non-epicenter area) | Assess the change in healthcare delivery because of the pandemic to patients with IBD from Hubei province (the epicenter of COVID-19) and Guangdong province (a non-epicenter area) | Front Med (Lausanne) | China | Not reported | No conflict of interest |
| **Lovecchio et al, 2021** | Provider confidence in TM spine evaluation: results from a global study | Assess the overall satisfaction and confidence in TM and the determinants of provider confidence | European Spine Journal | Multiple countries including in Africa, Asia Pacific, South America | No funding | No conflict of interest |
| **Gomes et al, 2020** | Impact of COVID-19 on clinical practice, income, health, and lifestyle behavior of Brazilian urologists | Assess the impact of covid on clinical practice, income, and lifestyle | International Brazil Journal of Urology | Brazil | Brazilian Society of Urology | No conflict of interest |
| **Li et al, 2020** | Pilot Study Using TM Video Consultation for Vascular Patients' Care During the COVID-19 Period | Assess the effectiveness and patient satisfaction of TM in vascular patients | Annals of Vascular Surgery | China | Not reported | No conflict of interest |
| **Ashry & Alsawy, 2020** | Doctor-patient distancing: an early experience of TM for postoperative neurosurgical care in the time of COVID-19 | Assess the effectiveness and safety of TM visits in providing postoperative care to neurosurgical patients | Egyptian Journal Neurology Psychiatry and Neurosurgery | Egypt | No funding | No conflict of interest |
| **Sultania et al, 2020** | Impact of the initial phase of COVID-19 pandemic on surgical oncology services at a tertiary care center in eastern India. | Assess the impact of the pandemic on the oncology services in the first 3 months since the first COVID case in the region | Journal of Surgical Oncology | Indonesia | Not reported | No conflict of interest |
| **Zhang, Yun et al, 2020** | Impact of COVID-19 outbreak on the care of patients with inflammatory bowel disease: a comparison before and after the outbreak in South China. | Assess the change in health care delivery and medication use before and after the COVID-19 outbreak | Journal Gastroenterol Hepatology | China | Not reported | No conflict of interest |
| **Zhang, Huang et al, 2020** | Application of remote follow-up via the WeChat platform for patients who underwent congenital cardiac surgery during the COVID-19 epidemic. | Evaluate the effect of WeChat based telehealth on post-operative follow-up for children who underwent congenital heart surgery during COVID | Journal of Cardiovascular Surgery | China | National Key Research and Development Program of China | No conflict of interest |
| **Lin et al, 2020** | Deep brain stimulation telemedicine programming during the COVID-19 pandemic: treatment of patients with psychiatric disorders. | Evaluate the effectiveness of post-operative care for DBS patients and the impact of the TM sessions | Journal of Neurosurgery | China | the Shanghai Clinical Research Center for Mental Health and SJTU Trans-med Awards Research | No conflict of interest |
| **Zhang, Zhu et al, 2020** | Utility of Deep Brain Stimulation TM for Patients with Movement Disorders During the COVID-19 Outbreak in China | Assess the utility of deep brain stimulation (DBS) TM in the management of patients with movement disorders | Neuromodulation Journal | China | no funding | No conflict of interest |
| **Zhang, Hu et al, 2020** | Implementation of a Novel Bluetooth Technology for Remote Deep Brain Stimulation Programming: The Pre- and Post-COVID-19 Beijing Experience | Assess the implementation of the Bluetooth technology for remote DBS | Movement Disorders Journal | China | the National Nature Science Foundation of China | No conflict of interest |
| **Tiwari et al, 2021** | Virtual Telephonic Follow-Up for Patients Undergone Septoplasty Amid the COVID Pandemic | Assess the feasibility of virtual telephonic consultation to the follow-up the patients in the immediate postoperative period | Indian Journal of Otolaryngology and Head & Neck Surgery | India | No funding | No conflict of interest |
| **Ferraris et al, 2020** | Recapitulating the Bayesian framework for neurosurgical outpatient care and a cost-benefit analysis of TM for socioeconomically disadvantaged patients in the Philippines during the pandemic. | Assess the responsiveness of TM to disadvantaged patients and the cost-benefit analysis of the remote intervention during the pandemic | Journal of Neurosurgery | Philippines | Not reported | No conflict of interest |

In total, 4,155 patients were included in this review. All studies included patients older than 18 who had experience with TM during the COVID-19 pandemic. Three studies reported patients’ education level, socioeconomic level, and distance from their home to the medical facility (Ferraris et al.,2020; Zhang et al., 2021; Li et al, 2020). Table 2 provides an overview of the study designs and the characteristics of the participants from the studies included. TM was implemented for a variety of purposes: remote treatment (3), education (1), general consultation (5), and post-operative follow-up (6). The most common TM application was post-operation monitoring and follow-up. TM was primarily delivered through phone calls (3) and video calls (13). Video consultations were conducted via platforms such as WeChat (4), online hospital platforms (3), treatment-related platforms (3), and other third-party virtual tools like HaoDaiFu (1) and Facebook Messenger (2).

**Table 2:** Study design of included studies.

| Author and Date | Research and/or survey design | Type of care | Key Findings | Setting (distance from medical facilities) | Limits of the study |
| --- | --- | --- | --- | --- | --- |
| **Qiu et al, 2020** | Electronic multiple-choice questionnaire about change of medications, procedures, and healthcare mode | physician consultation, not specified | More patients in epicenter than in non-epicenter had delayed procedures<br>Increased use of telemedicine after the outbreak compared to before the outbreak in epicenter, while such a significant increase was not observed in non-epicenter<br>2/3 from both sides agreed that TM should be used more in the future | N.R. | Non responsiveness to the survey by very sick patients can skew findings<br>The education level, the disease condition, the age, and the timing of the survey can bias the results |
| **Lovecchio et al, 2021** | Anonymous survey Delphi approach with 4 rounds of a total of 42 questions, collecting participant's experience with telemedicine and in comparison, with in-person visits | pre-operative for triage, post-operative care | 57.5% used videoconferencing platforms that are EMR-integrated or not<br>34.6% used phone calls<br>No statistical difference in confidence in telemedicine between regions<br>Videocall considered higher than audio calls.<br>No statistical difference in specialty in confidence<br>74.9% of physicians trust TM<br>TM inferior the physical patient evaluation. | N.R. | Response to items optional for the survey<br>The timing of the survey (during COVID-19) can influence the attitude, in need for an update of opinions post-pandemic<br>Study not controlled to rule out associations<br>No explanation for differences between regions<br>The survey available for only two weeks, low participation rate (12.5%)<br>Participants mainly from urban areas |
| **Gomes et al, 2020** | A 39-question, web-based survey (using SurveyMonkey) to analyze changes in clinical practice and income | physician consultation, not specified | Reduction of 83.2% of patient visits, 89.6% of elective and 54.8% emergency surgeries<br>Income reduction by 54.3%<br>38.7% . used video consultations<br>49.2% confident with >80% their TM<br>62.7% of patients accepted TM and satisfied | 45% respondents located in capital state<br>61% work in their private clinic, 14.1% in private sector and 14.6% in public sector | Participants representing only 18.2 % of the invited pool<br>No evaluation done between differences in usage of TM in public and private sector. |
| **Li et al, 2020** | Questionnaire on patient's satisfaction at the end of each TM episode, scoring option of 1-5 | post-operative and virtual consultation (not specified) | Mean time of the call was 11±8.9 minutes-> short consultation<br>95% preferred remote telemedicine rather than postponing.<br>48% of patients were with family members during the call.<br>Advantages of telemedicine: personal (95%) and cheaper (97%)<br>90% lack of physical contact is not a problem. One patient (1%) had worsening condition leading to seek urgent care | 25 patients were outside of Hubei, the province of the facility, when receiving the call.<br>Distance from home to hospital 324 ± 678 km.<br>Average driving time of 194 mins at a speed of 100 km/h | 69% of those contacted participated descriptive study without analysis on the advantages/disadvantages<br>Lack of evidence that TM can replace in person consultation<br>Cohort of patients heterogeneous, with different comorbidities and medical priorities |
| **Ashry & Alsawy, 2020** | Satisfaction survey to patient (6 questions) and physician (5 questions) with Yes/No option, provided at the end of each TM session | Post-operative | Total number of TM visits: 67 visits (average 2.23 visits for each patient).<br>Total number of emergency calls: 32 calls (average 1.06 calls from each patient).<br>Patient/doctor satisfaction: 90%, prefer TM in the future (90/95), remote examination (80/80) satisfied with audio and visual application (80/90).<br>Patients (100%) saved time and money<br>Female patients: refusing TM due to lack of privacy<br>Inability to perform accurate remote physical clinical examinations<br>Literacy necessary for TM usage<br>20% of doctors reported difficulties in performing remote clinical evaluation | Most patients needed > 4h to reach the hospital.<br>Distance to hospital:<br>3 patients < 1 hours<br>7 need 1-4h<br>20 need > 4 hours | Results not representative of the whole population:<br>40 refused to participate in this study and preferred in-person visits and opinion not included<br>12 excluded, did not have smartphones<br>8 patients had problem hearing, visual<br>20 were illiterate |
| **Sultania et al, 2020** | Hospital database |  | Total 261 teleconsultations, with 676 unique patients<br>Residents divided into two groups to avoid exposure and functioned alternate weekly with one group handling the OPD patients and teleconsultations for a week and the other focus on wards<br>Low COVID-19 infection within the facility<br>After the lockdown was over, the in-person visits and admissions and surgery started increasing | N.R. | No evaluation done between differences in utility for TM vs in-person.<br>The increase of in-person visits post-lockdown was not analyzed.<br>No control groups. |
| **Zhang, Yun et al, 2020** | Electronic questionnaire surveys on gastroenterologists and IBD patients' attitudes towards TM | | In post-outbreak period, 64.98% physicians have used TM with an increase compared with 46.13% in the pre-outbreak period.<br>65.54% supported TM use in the future<br>Compliance to medication the same pre/post pandemic.<br>Utility for each platform: WeChat (88, 64.23%), third-party online clinic such as HaoDaiFu (81, 59.12%), and online clinic from their hospitals (38, 27.74%)<br>Significant difference between older ($\geq 40$ years) and younger patients ($< 40$ years) on interest in TM: 71.2% vs 51.4%<br>59.6% patients hoped that more doctors will have TM<br>42.38% want more improvement in insurance coverage and convenient drug delivery. | N.R. | Retrospective of a single-center, not representative of all centers<br>Need comparison between IBD centers<br>No description of the different components within the TM platform<br>No comparison between the different types of TM |
| **Zhang, Huang et al, 2020** | Analysis of medical and family data of pediatric patients who used TM for post-surgery follow-up as well as self-reported scales (SDS and SAS) for psychological evaluation | post-operative care, educative platform, and psychology care to parents | Little knowledge on the disease by parents who were unable to fully understand the information provided during hospitalization.<br>57.4% (62/108) of the parents have high school degree or lower.<br>2 children with poor incision were healed after WeChat TM session<br>Nutrition guidance prevented heart failure in children<br>A high amount of stress anxiety and depression in the parents of the patient | 76% live in rural area:<br>Complete blockage of interprovincial travel<br>public transport limited<br>All advanced medical care in cities | A single-center retrospective study<br>small sample.<br>No control groups<br>Timing of the study, not representative of all different timing<br>No correlation done on the parents' psychological status and the clinical outcomes reported |
| **Lin et al, 2020** | Analysis of clinical data of 4 selected patients and their TM sessions | treatment | No unexpected loss of connection during the sessions.<br>Psychological support and troubleshooting provided during the TM sessions<br>Modest to substantial clinical improvements after DBS surgery were observed in some but not all patients (compliance)<br>Stimulation-related side effects were reported by 2 patients and included reversible sleep and mood problems, headache, and a sensation of heat | N.R. | DBS telemedicine management for movement disorders is related to internet connection issues, such as video transmission delay, which might considerably affect the rating of visually assessed motor items (e.g., finger tapping). |
| **Zhang, Zhu et al, 2020** | 40 hospitals database on participant and their DBS treatments and survey on satisfaction with their TM | treatment | Monthly number of DBS telemedicine sessions requested by patients higher during the COVID-19<br>High patient satisfaction rates<br>Increase number of patients requested their first post-operative parameter adjustments through DBS tele-programming, which did not happen in 2019.<br>A greater need for DBS telemedicine during the COVID-19 outbreak, but also patients and physicians may had become more familiar<br>High number reflect both an increased familiarity and acceptance of this novel technique and a greater need due to the COVID-19 outbreak and implementation of protective measures. | N.R. | No comparison between the clinical effectiveness of DBS TM and in-person DBS treatment using standardized clinical rating scales.<br>The selected hospitals all used the same DBS telemedicine system therefore; it is unknown if the results can be generalized to other hospitals using other DBS TM systems. the study period covered the main, and not the entire, period of the COVID-19 outbreaks in China. We intend to report data obtained before, during, and after the COVID-19 outbreak in a future publication, which will provide a more comprehensive picture of the role and value of DBS telemedicine for outpatients with movement disorders in public health emergencies. |
| **Zhang, Hu et al, 2020** | Analysis of patient's hospital data and their TM sessions | treatment | 402 patients had their DBS adjustment sessions<br>386 cases of medical optimization<br>Tele-programming adjustment led to 25 cases of dysarthria and 38 patients of dizziness. These were identified and quickly managed through the remote program system. => timely and efficient and quick. | Mean travel distance: 1141km ± 825 km | Lack information of methodology, inclusion and exclusion criteria provided |
| **Tiwari et al, 2021** | Satisfaction questionnaire and NOSE score (Nasal obstruction and symptom evaluation) | post-operative | mean duration of virtual follow up: 36.7 days<br>All patients had improvement in symptoms with 14 patients completely asymptomatic<br>83% highly satisfied by the telephonic follow-up and accepted that telephonic follow-up was convenient for them.<br>Less post-operative complications observed | NR | The time coverage not reflective of the whole period of the Pandemic.<br>Very small sample<br>No comparison to a control or in-person visits |
| **Ferraris et al, 2020** | Patient survey on TM (questions with ordinal choices) and expenditure patterns (open-ended questions) in both English and the Pilipino | General, post-operative follow-up | High rates of utility for the patients and their caregivers<br>64% of respondents: the affordability of TM preventing them from further impoverishment<br>The benefit-cost ratio was 3.51<br>Adverse life events and financial risk averted by TM<br>TM reduces the gap in access to specialty care<br>Score ranging from (8-10)/10 for efficiency (91%), adequacy (87%), timeliness (78%), satisfaction (89%).<br>66.7% participants from low-income groups | 77.4% live far from the center | Only 57 out of 272 responded to the survey.<br>Patients with heterogeneous clinical profiles<br>the limited study duration<br>Confounding factors not accounted<br>No evaluation of the clinical efficacy of each kind of care through TM |

The main themes discussed across the studies are summarized in table 3. Five studies focused on patients’ perceived value of TM and satisfaction rates. Four focus on the physician’s perspective, their perceived value of TM, and their satisfaction rate. One mentioned the utility of TM for education and mental care for caregivers. Three examined TM’s role in remote treatment post-implant surgery.

**Table 3:** Concepts identified in included studies.

| Concepts |  | Qiu et al, 2020 | Lovecchio et al, 2021 | Gomes et al, 2020 | Li et al, 2020 | Ashry & Alsawy, 2020 | Sultania et al, 2020 | Zhang, Yun et al, 2020 | Zhang, Huang et al, 2020 | Lin et al, 2020 | Zhang, Zhu et al, 2020 | Zhang, Hu et al, 2020 | Tiwari et al, 2021 | Ferraris et al, 2020 |
| --- | --- | --- | --- | --- | --- | --- | --- | --- | --- | --- | --- | --- | --- | --- |
| Patient perceived value | Satisfaction rate |  |  | ? | ? | ? |  |  |  |  | ? |  | ? | ? |
|  | Preference for TM over in-person |  |  |  | ? |  |  |  |  |  |  |  |  | ? |
|  | Affordability/ cost-effectivity |  |  |  | ? | ? |  |  | ? |  |  |  | ? | ? |
|  | Time saving/ less travel |  |  |  | ? | ? | ? |  | ? |  |  |  | ? | ? |
|  | Interest in continuing TM in the future | ? |  |  | ? | ? |  | ? | ? |  | ? |  | ? | ? |
|  | Personalized |  |  |  | ? |  |  |  | ? | ? | ? | ? |  |  |
|  | Safety |  |  |  |  |  |  | ? | ? | ? | ? | ? |  |  |
|  | Increased accessibility | ? |  |  | ? |  |  | ? | ? | ? | ? | ? | ? | ? |
| Provider perceived value | Satisfaction rate |  | ? | ? |  | ? |  | ? |  |  |  |  |  |  |
|  | Confidence in clinical outcome |  | ? | ? |  |  |  |  |  |  |  |  |  |  |
| Effectiveness of TM | Equivalence to in-person consultation |  | ? | ? |  |  |  |  |  |  |  |  |  |  |
|  | Quantity of info (amount of TM or time spent/patient) |  |  |  | ? | ? | ? | ? | ? |  | ? |  | ? |  |
|  | Accuracy |  | ? |  |  |  |  |  | ? | ? | ? | ? |  |  |
|  | Reduce burden on health system |  |  |  |  | ? | ? |  | ? |  |  |  |  |  |
|  | Improved clinical outcomes |  |  |  |  |  |  |  | ? | ? | ? | ? |  |  |
| Feasibility of TM | Consistency and standards |  |  |  |  |  |  |  |  |  |  | ? |  |  |
|  | Timely |  |  |  | ? | ? |  |  | ? | ? | ? | ? |  |  |
|  | Flexibility in time or type of platforms | ? |  |  | ? | ? |  |  | ? | ? |  |  | ? | ? |
|  | Presence of reimbursement/ regulation |  |  | ? |  |  |  | ? |  |  |  | ? |  |  |
| System Design of TM | Connectivity |  |  |  | ? | ? |  |  |  | ? | ? |  |  |  |
|  | Security |  |  |  |  | ? |  | ? |  |  |  |  |  |  |
|  | Privacy |  |  |  |  | ? |  |  |  |  |  |  |  |  |
| Limits of TM | Not equivalent for physical exams |  | ? |  | ? | ? |  |  |  |  |  |  |  |  |

### c. Synthesis of results

#### The perceived value of TM

**1. Satisfaction**: All studies included indicated that TM is a valuable tool for clinical diagnosis and patient monitoring. Six studies reported that most patients perceived TM from satisfactory to very satisfactory across different platforms [15–18]. High rates of utility were recorded by caregivers and patients. A study conducted in the Philippines reported high scores for perceived efficiency (91%), adequacy (87%), timeliness (78%) and satisfaction (89%) [19]. 83% of patients were also satisfied with the telephonic follow-up and indicated being willing to use TM in the future (Tiwari et al., 2020) with greater interest in patients aged 40 and older compared to younger ones (Ashry & Alsawy, 2020). The care provided through TM was described by patients as personalized and safe (Li et al., 2020). Regarding physician’s general satisfaction, it is slightly higher compared to patients, while the difference is not statistically significant (Ashry & Alsawy, 2020). Overall, a high satisfaction rate was observed across specialties and settings.
**2. Access to care**: TM increased access to care with its cost-effectiveness and affordability, the reduced time commitment, and flexibility. The Ferraris et al., 2020 study found the benefit-cost ratio at 3.51, indicating that the benefits of TM outweighed the costs. TM also improved access to specialized care such as neurosurgical care. An increase in request for TM sessions for Deep Brain Stimulation treatment (DBS) for follow-ups was also observed [20]. Therefore, TM facilitated access to care and increased timely use of care services.
**3. Time commitment to care:** TM reduce the. time travel to get care, thus removing the geographical barrier that was expressed by many patients in the surveys and across the different countries. In the study from the Philippines, 77.4% live far from the center (Ferraris et al., 2020). The mean travel distance to Beijing for DBS treatment was 1.141 km (Zhang, Hu et al., 2021). The distance from home to the hospital was indicated to be 324km for patients in Hubei and the estimated average driving time would be 194 minutes at a speed of 100 kilometers per hour (Li et al, 2020). Most patients required more than 4 hours to reach the hospital in Egypt (Ashry & Alsawy, 2020). Pandemic-induced mobility restrictions, including suspension of public transportation and interprovincial travel bans, further exacerbated geographic barriers to surgical access (Zhang, Huang et al, 2020). Some had to spend one night near the facility due to the travel restriction curfew (Ashry & Alsawy, 2020).
**4. Affordability:** According to Ferraris et al., 2020, 64% of the respondents have indicated that the affordability of TM prevented them from further impoverishment as they won’t need to borrow money from others. With TM, several adverse life events and financial risks were averted such as borrowing money, seeking a babysitter, losing income, discontinuation of children’s education, being forced to accept charity and pawning their possessions [19]. TM can serve as financial protection for the low-income population and improve access to specialty care. Another example is families with increased unfavorable economic conditions due to COVID-19 still receive care with TM in India (Ashry & Alsawy, 2020).

#### Effectiveness of TM

**1. Clinical outcomes**: Four studies mentioned improvements in clinical outcomes. Remote Deep Brain Stimulation (DBS) was associated with sustained to modest improvements in psychiatric disorders (Lin et al., 2020) as well as timely adjustments in motor disorders (Zhang, Zhu et al., 2020). Less post-operative complications were observed using TM for follow-up (Tiwari et al, 2021). Notably, TM helped reduced COVID-19 transmission in the community, reducing the infection rate for physicians and patients with a weak immunity [21]. However, these results are limited, with the absence of control groups. More research with a control group is needed to determine their standardized clinical benefit.
**2. Physicians’ confidence rate in their diagnosis:** Only two studies (Lovecchio et al., 2021 and Gomes et al., 2020) focused on the physician’s level of confidence. 49.2% of urologists have been indicated to be confident in more than 80% of their consultations (Gomes et al., 2020). 64.3% of spine surgeons indicated that TM is equivalent to in-person visits regarding taking patients history and 72.3% believe that it is TM equivalent to reviewing, and explaining imaging (Lovecchio et al., 2021). They also stated that they feel more confident in their ability to formulate and communicate a treatment plan through TM compared to in-person visit imaging (Lovecchio et al., 2021).
**3. Platforms for TM:** Both phone calls and video calls were reported to have a positive outcome and high satisfaction rate. One study compared two types of TM platforms. The use of a video-call platform was shown to have increased confidence in the diagnosis formulation by the physician compared with audio calls only (Lovecchio et al., 2020). Video calls were conducted either through hospital-affiliated TM platforms or existing social media like WeChat and Facebook Messenger. While some studies provided different platforms of TM, only general satisfaction was reported without specific platform analysis.

#### Role of TM

**1. TM for treatment:** Three studies evaluated the use of TM for Deep Brain Stimulation (DBS) [20, 22, 23]. All of them reported that remote DBS treatment was adequate and capable of proper parameter adjustments for the voltage, frequency, pulse width, and contact site [20], and effective optimization of medication [22]. TM could effectively detect and diagnose medical conditions that needed immediate attention or prescription update. In the Ashry et al., 2020 study, 3 wound infections were detected, 10 reported uncontrolled pain resulted in the prescription of pain killers, and 2 focal seizures led to change in antiepileptic drugs. No connection or software-related malfunction has occurred during the TM sessions, making TM intervention reliable. No severe medical adverse effects were reported as well, thus proven to be safe. Some stimulation-related side effects were reported but were reversible [20]. The sleep, mood, headache, and heat problems detected in the Lin et al., study were reported and reversible after re-adjustments. In the first three months of 2020, patients are more willing to have their first DBS post-operative adjustment session through TM than in 2019 [20].
**2. TM for follow-up and monitoring:** TM in general surgery and other surgery specialties is mainly used for post-operative follow-up and monitoring. Consistent results included reliable monitoring tool and reduction of complications (Tiwari et al, 2021). Side effects were detected and resolved promptly, preventing surgical infections (Zhang, Huang et al, 2020). Post-operative surveillance is a challenging task for health systems. TM has the capacity to relieve the management burden of post-discharge patients on hospitals by reducing the in-person visits.

#### Challenges in feasibility and acceptance

**1. Feasibility:** The remote nature of TM allows easy access to care. For example, an emergency number is provided and can be used aside from TM video calls (Ashry & Alsawy, 2020; Ferraris et al, 2020). However, this led to over-utilization when it was not necessary (Ashry & Alsawy, 2020). One reported challenge in feasibility was the use of TM by the elderly (Ashry & Alsawy, 2020). This limit is mitigated with the attendance of family members at the virtual consultation. Due to their lowered mobility and higher risk of morbidity, this group would, nevertheless, also stand to gain the most. It was also recorded that poor network connections and new TM platforms made the use confusing to some (Ashry & Alsawy, 2020). To make sure that patients can use the platforms easily, simulation sessions were provided before their discharge (Ashry & Alsawy, 2020).
**2. Limitations in physical examinations:** One limit of TM mentioned is their inability to replicate the hands-on physical exams and will probably continue to be their largest limitation (Lovecchio et al., 2021). Around 90% of spine surgeons reported that TM was slightly worse than in-person visits for these tasks (Lovecchio et al., 2021). One study cited that some surgeons were unable to perform accurate remote clinical examinations such as motor power assessment in some spinal cases. More research is needed for modern TM that can perform higher quality remote examinations (Ashry & Alsawy, 2020).
**3. Acceptance:** The acceptance for TM by patients is consistently high across the studies that mentioned it. 62.7% of patients accepted TM as an alternative and are satisfied (Gomes et al, 2020). In Egypt, high rates of illiteracy and limited familiarity with technology was reported which created some resistance in both physicians and patients. Some physicians are accustomed to traditional way to diagnose, and a large percentage of women do not feel secure during online consultations due to a lack of privacy (Ashry & Alsawy, 2020). Therefore, an education initiative is needed to improve awareness of TM users about their importance, purpose, and benefits. Proper TM training should be added to medical education as well. More guidelines and regulations on patients’ data confidentially will also help improve acceptance.
**4. Limited infrastructure:** TM is dependent on the existing telecommunication and is the major aspect for continuing innovation. Two main challenges were reported: the lack of adequate medical-focused virtual platforms, and network services. Due to the limited TM options in some countries, many have opted to use every day social media as their virtual TM. However, the use of such platforms such as Facebook Messenger for medical purposes, questions the confidentiality, safety, and security of such intervention (Ashry & Alsawy, 2020; Ferraris et al, 2020). Some others like WeChat, created an additional medical layer to their app dedicated to TM (Qiu et al, 2020; Li et al, 2020; Zhang, Yun et al, 2020). One study recorded that 20% of patients were not satisfied with their TM visits due to poor internet connections (Ashry & Alsawy, 2020). While there were some connection problems, the satisfaction was overall high, and TM allowed the provision of care to remote areas during lockdowns.
**5. Lack of regulation/guidelines:** The absence or limited guidelines has been a challenge for both providers and patients, increasing resistance in both users. Very few studies indicated the authorization or regulation of their TM intervention. Thus, questions can be raised regarding security and safety of TM platforms. In Egypt, there is a lack of publications on TM implementations leading to a gap in knowledge on what should be done regarding security standards and patients’ confidential data (Ashry & Alsawy, 2020). Little is also known regarding reimbursement, a problem raised in the Gomes et al., 2020 paper. Only 49.5% of urologists charged more than 80% of their video consultations. Billing regulation was currently not fully established, and TM is only temporarily authorized (Gomes et al., 2020). In the study led by Zhang, Yun et al., 2020, 42.38% of patients and 75.6% of physicians believed that TM coverage by insurance should be increased. In China, TM was formally recognized with approvals by the National Medical Products Administration, for tele-treatments such as remote DBS (Zhang, Hu et al, 2020). Therefore, the gap in policy was highlighted in several papers, indicating an important need for transparency and regulation.

## 4. Discussions

### a. Summary of evidence

COVID-19 disrupted surgical services in LMICs, leading to delayed lab procedures, postponed elective surgeries, and intravenous transfusion [24]. Reduction in care services was observed by 50% in the non-epicenter, 80% in the epicenter, and “no show” increased twice in both locations [24]. Perioperative and post-operative patients are particularly vulnerable individuals, especially those with high relapsing and remitting conditions such as IBD. TM can alleviate administrative burden in health monitoring and provide timely therapy adjustments for preventable admissions. Evidence suggests that TM is an efficient, cost-effective tool to treat patients, particularly when considering patients’ travel time and cost. It enhanced access to care, and provides positive clinical outcomes, and timely and accurate treatments remotely. Telemedicine was associated with increased provider confidence, with most clinicians perceiving it as a viable substitute to in-person consultations. Although telecommunication and connectivity infrastructure limitations were reported, particularly in rural areas, TM use remained high due to the widespread ownership of smartphones.

The mean age of all participants is 46 years old, limiting generalizability to younger populations. Patients older than 40 years old are more significantly interested in TM than patients younger than (< 40 years). More than half of the participants older than 40 want more doctors to adopt TM (Zhang, Yun et al., 2020),

This synthesis also indicated that while the population overall is interested in using TM in the future, the interest is higher in rural areas, where the income is lower and the distance to health facilities is more important (Ferraris et al., 2020). COVID-19 exacerbates financial resources, restricted travel, and reduced public transportation. Urban areas suffer less from these consequences as most health facilities are in the city. Therefore, TM may serve as a structural equalizer by attenuating longstanding geographic and financial barriers to surgical care in underserved LMIC populations.

### b. Expansion and collaboration with other specialties

In the Lin et al., 2020 study, the idea of expanding current TM was provided by combining existing remote DBS treatment with psychological counseling, medical consultations, and medical prescription to improve adherence and reduce poor symptom control observed in patients. In addition to the regular pediatric post-surgery TM monitoring, an educative platform on nutrition and psychological support care was provided to the parents which successfully prevented heart failure in participating children and taking care of their caregivers’ mental health (Zhang, Qi-Liang et al., 2020). Further piloting is warranted to evaluate such collaboration for monitoring and prevention.

### c. Stakeholders’ analysis

A stakeholders analysis reveals the diverse motivations, interests, and level of influence [25]:

#### Patient/individual users

They have expressed high satisfaction and interest in TM during the pandemic but also for future use. TM was shown to provide flexible and real-time care, that can prevent severe side effects. Populations living in remote areas and far from health facilities are most interested (Qiu et al, 2020). However, patients have limited agency or influence over implementation decisions.

#### Care Providers

Physicians indicated to be satisfied and confident with TM because it serves as an adequate and efficient monitoring tool. However, increase in non-urgent calls was reported, due to the feasibility nature of TM, straining the existing workforce (Ashry & Alsawy, 2020). Physician’s interest in TM is driven by patient demand and the opportunity for a new source of income. More transparency in terms of reimbursement and use is needed (Gomes et al., 2020). Countries such as Brazil only temporarily allowed TM, therefore, the ambiguity in regulatory makes it less attractive to providers. Physician interest in TM remains moderate due to regulatory uncertainty and workload concerns.

#### Hospitals

Most of the studies included were investigated in public and/or academic medical hospitals, thus there is an emphasis on improving care access. Kasr Alainy Hospital and the Jose R. Reyes Memorial Medical Center, for example, are health facilities that primarily focus on the low-income and socioeconomically disadvantaged population. However, the lack of any established guidelines prompted local facilities to form their management plan (Sultana et al., 2020). To conclude, the interest can depend on the population that the facility mainly serves, with little power in policymaking. However, hospitals can influence policy decisions through their research findings.

#### Governments

While TM is attractive because it can reduce the infection rate and potentially decrease healthcare expenditure due to its cost-effectiveness, the lack of existing infrastructures, guidelines, and e-readiness indicates that more financial investment should be done. Limited knowledge TM’s long-term impact makes it less attractive to governments. Studies suggest a lack of interest for long-term TM implementations (Gomes et al., 2020). While policy influence is high, the interest can be categorized as low and short-term.

### d. Limitations and policy recommendations

#### 1. High risk of bias

The availability of studies on TM in surgery in LMICs is limited, reflective of the rapidly changing clinical practice and uncertainty. The heterogeneity of the included studies prevented any method of meta-analysis; thus, no statistical power can be drawn. Language restrictions may have excluded relevant studies published in non-English languages. The lack of randomization in the study design indicates that the participants sample was not adequately representative of the target population. The heterogeneity within the studies regarding the medical conditions and the demographics of the participants limit generalisability. Finally, the unequal representation of studies from Asia is not representative of the overall situation in LMICs. Future research should incorporate a broader representation of LMIC settings and surgical sub-specialties to enhance generalisability and comparative validity.

#### 2. The satisfaction rate as an outcome is limited

Satisfaction rate as an outcome is limited due to the use of different instruments and scales across the studies. While patient satisfaction has been commonly chosen to indicate the quality of care, their overall reported satisfaction might be related more to their expectations, thus making it a more subjective criterion [26]. Literature evidence indicated that satisfaction rate disposes of a ceiling effect, which means that the measure will not increase higher after a certain threshold is reached. Therefore, the findings do not fully illustrate the full spectrum of satisfaction and do not reflect the clinical outcome. Although we attempted to identify the different dimensions in included studies, further research should be conducted following the conceptual framework below (Figure 3).

**Figure 1:**
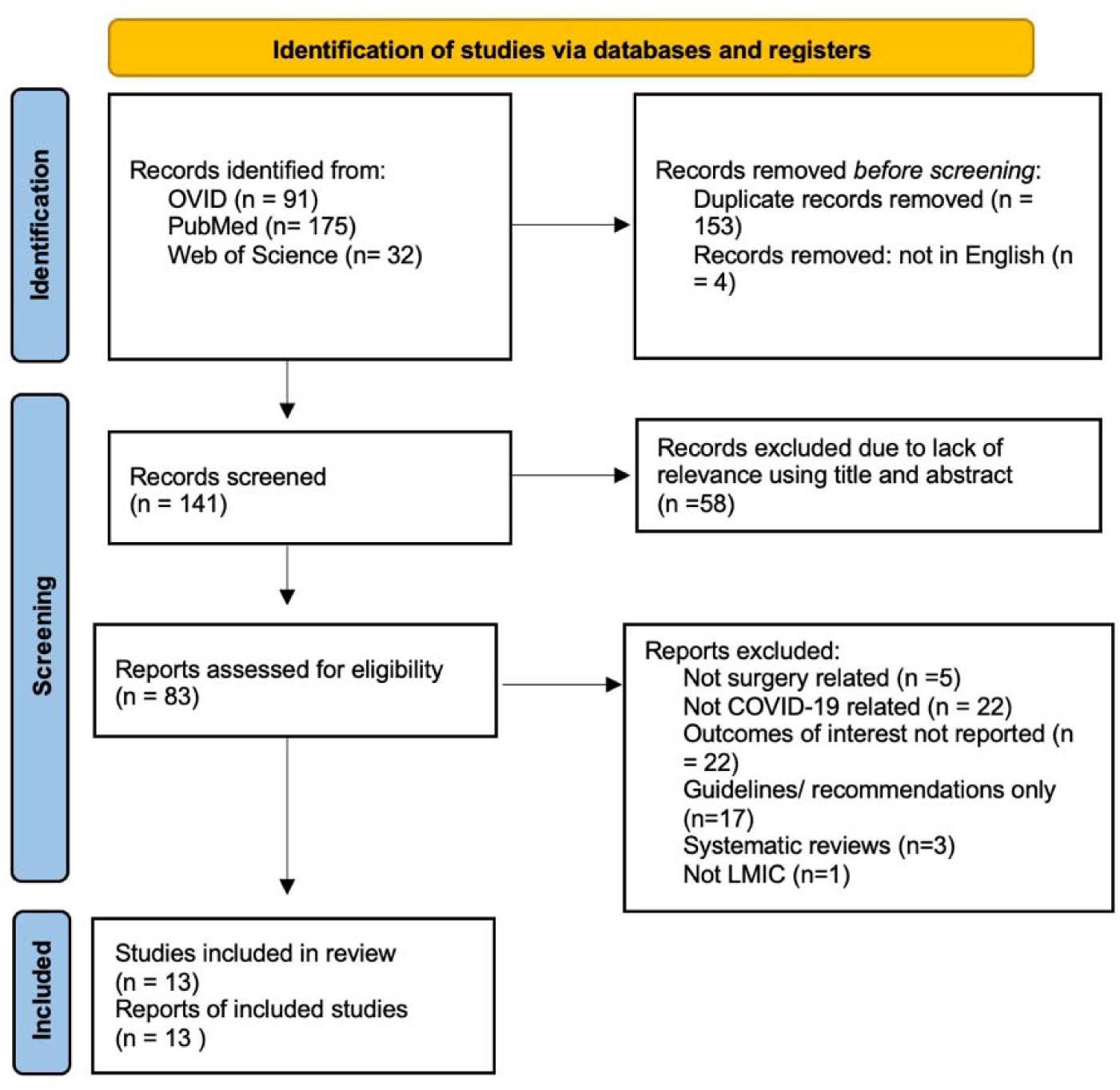
Flowchart of the PRISMA diagram with the search strategy and selection.

**Figure 2:**
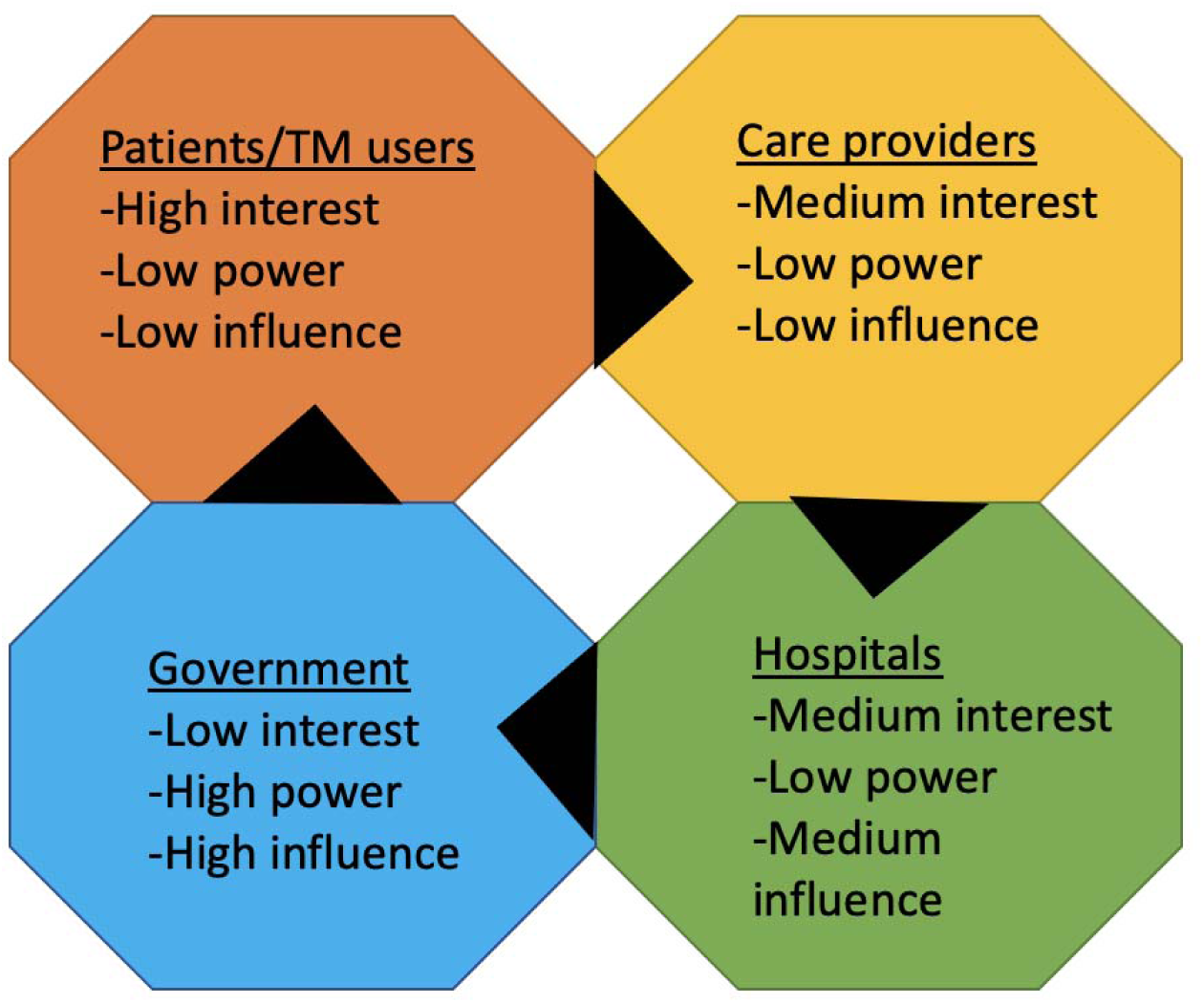
Stakeholders’ stand on the implementation of TM

**Figure 3:**
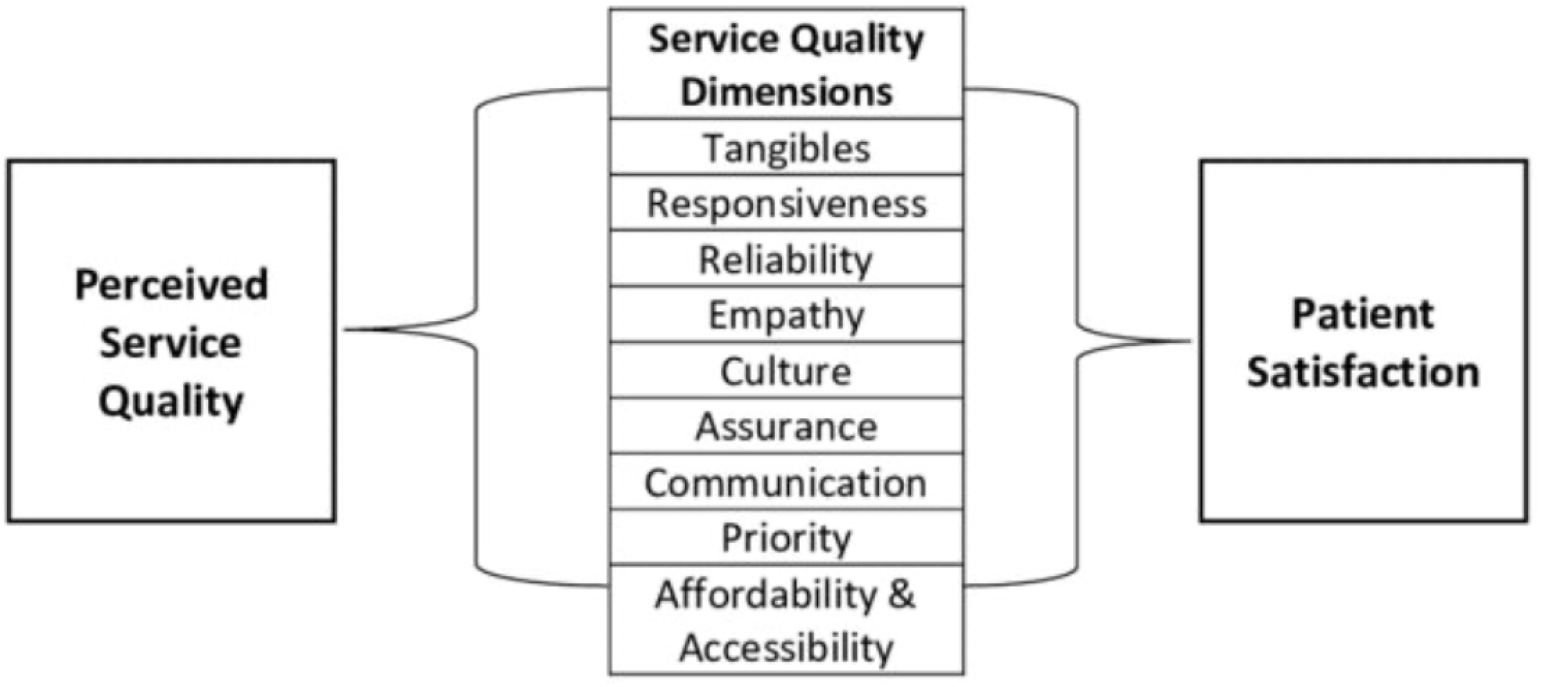
Conceptual framework of the multi-dimension of satisfaction[27].

**Figure 4:**
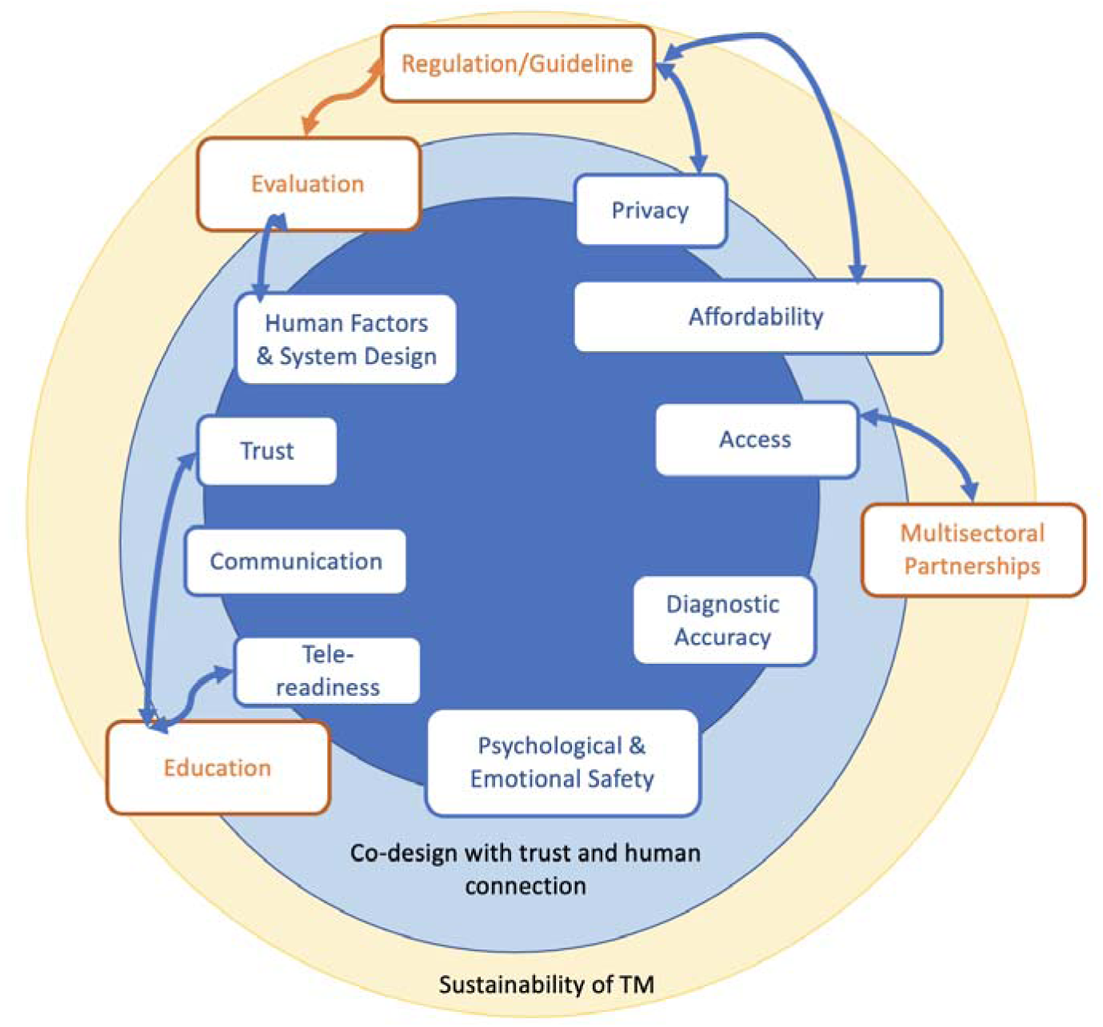
Proposal for conceptual framework of patient-centered TM for LMIC settings.

#### 3. Lack of safety and security-related outcomes

The lack of rigorous safety data questions the clinical quality of TM. While some side-effects of severe complications were identified using TM, there is a lack of input to determine if TM was preventing them similarly to an in-person visit [17]. The same can be said regarding the security aspect of TM, with the use of everyday online platforms such as Facebook Messenger to share sensitive and personal information. Due to the emergency context elicited by the COVID-19 Pandemic, authorization and guidelines for TM are temporary for some countries like Brazil. Recurrent evaluation mechanisms are needed to determine TM’s clinical outcomes and to regulate medical records protection.

#### 4. Beyond the pandemic, towards a sustainable and resilient healthcare

While TM cannot yet replace in-person visits, it improves access to care, alleviates some burden from health systems and provide an alternative source for flexible and timely medical attention. Considering the gap in access to care pre-pandemic and the current impact of TM on health systems in LMICs, a move towards sustainable TM can have positive lasting consequences.

An updated conceptual framework of TM was proposed to consider the necessities for a successful implementation in an LMIC setting. Tele-readiness, affordability, trust, and functionality emerged as key factors in patients’ and providers’ acceptance. They should therefore be added to the existing framework. Policymakers should implement measures focusing on these aspects according to the country’s current concerns and priorities. Multisectoral partnerships can be formed to improve the existing telecommunication infrastructures, increasing network access to the rural areas, which is the population most interested in TM usage. Improved tele-readiness can enhance trust. The awareness and understanding of the benefits of TM needs to be made available to the population and healthcare providers, reducing the resistance to practice change. The move from temporary to complete authorization of TM can initiate the standardization of care with TM and improve the affordability to care services. Finally, trust and acceptance can be increased with careful evaluation and update of the existing design of TM to integrate the preferences and the needs of the patients and providers.

Drawing from the evidence synthesis and conceptual framework, policy recommendations included an interoperable and user-friendly TM platform with integrated EMR. Pilot programmes can be implemented to understand the population’s interests and needs. A seamless integration of different TM services for continuity of care, preventing loss of patients across the care pathways. Establishment of reimbursements for TM to incentivize. Confidentiality and privacy should be guaranteed across the three stages of TM.

## 5. Conclusion

In efforts to reduce post-COVID surgical access problems, physicians and health systems across the globe have implemented TM in their care delivery. While TM was rapidly implemented by COVID-19, it disposes of numerous promising outcomes, such as reduction of adverse effects post-surgery and timely care access. While high satisfaction and confidence rates were reported in both patients and physicians, challenges in replicability of in-person visits, reliable network connection, and regulations make TM less attractive and accepted by many. Improving the current TM infrastructure is essential to address persistent access barriers in LMICs. The next step includes enhancing its value by standardizing virtual examinations with evidence-based guidelines and regulations. Further research is required to determine TM’s specific utility to align implementation and the existing needs in surgical care in LMICs.

## Data Availability

All data produced in the present work are contained in the manuscript

## Appendix 1 Search Strategy

### Embase (OVID)

1. ’telemedicine’/exp OR ‘telehealth’/exp OR telemedicine:ti,ab OR telehealth:ti,ab OR e-health:ti,ab OR mHealth:ti,ab OR ‘remote consultation’:ti,ab
2. ’surgery’/exp OR ‘surgical procedure’/exp OR surgery:ti,ab OR surgical:ti,ab OR operation*:ti,ab
3. ’covid-19’/exp OR covid-19:ti,ab OR coronavirus:ti,ab OR sars-cov-2:ti,ab OR pandemic:ti,ab
4. ’developing country’/exp OR lmic:ti,ab OR “low and middle income countr*“:ti,ab
5. 1 AND 2 AND 3 AND 4
6. NOT ‘systematic review’/exp
7. Limit 6 to (English language and human)

### Pubmed

((“Telemedicine“[Mesh] OR “Telehealth“[Mesh] OR “Remote Consultation“[Mesh]

OR telemedicine OR telehealth OR mHealth OR e-health OR “remote consultation”)

AND

(“Surgical Procedures, Operative“[Mesh] OR surgery OR surgical OR operation)

AND

(“COVID-19“[Mesh] OR COVID-19 OR coronavirus OR SARS-CoV-2 OR pandemic)

AND

(“Developing Countries“[Mesh] OR LMIC OR “low- and middle-income countries”))

NOT “systematic review“[Publication Type]

Filters: Publication date from 2019/12/31 to 2022/07/28; English; Humans

### Web of Science

TS= (telemedicine OR telehealth OR “remote consultation” OR e-health OR mHealth)

AND

TS=(surgery OR “surgical care” OR operation*)

AND

TS=(COVID-19 OR coronavirus OR SARS-CoV-2 OR pandemic)

AND

TS=(“low and middle income countr*” OR LMIC OR [insert World Bank LMIC country names])

NOT TS=(“systematic review”)

Refined by: Languages (English) AND Document types (Article)

Timespan: 2019-2022

